# “Impact of sociodemographic factors on the relationship between perceived general health and number of healthcare visits”

**DOI:** 10.1101/2023.06.18.23291552

**Authors:** Shirin Saleh, Jazmine Abril

## Abstract

This retrospective cross-sectional study examines the relationship between the number of healthcare visits, perceived health status, and sociodemographic characteristics. Using data from the National Health and Nutrition Examination Survey (NHANES) for 2017-2018, the study analyzed responses from 4,755 participants aged 20-80. Logistic regression analysis was used to assess the association between healthcare visits and perceived health status while controlling for sociodemographic factors. The results revealed that increased healthcare visits were significantly associated with a lower likelihood of reporting good or better health status. This association remained significant even after adjusting for sociodemographic characteristics. However, gender was not found to be a significant predictor of perceived health status. Other sociodemographic factors, including age, race, education, marital status, and annual household income, were all significant predictors in the model.

Furthermore, the study identified racial disparities in perceived health, with Hispanic and Non-Hispanic Black individuals reporting lower rates of good or better health than Non-Hispanic White individuals. Higher levels of education were associated with better-perceived health, highlighting the importance of health literacy in healthcare access. Additionally, income was found to mediate the relationship between healthcare visits and perceived health status, with individuals of higher income more likely to perceive their health as better.

## Introduction

Perceived health status refers to an individual’s subjective evaluation of their overall health. Many health surveys conducted among countries belonging to the Organization for Economic Co-operation and Development (OECD) incorporate questions about perceived health status to gather individuals’ assessments of their general well-being (OECD, 2021).

Studies have shown that perceived health among older adults is associated with socioeconomic factors (age, gender, income, and education), mental status (cognitive impairment, depression), and physical function in general (Machón et al., 2016; Shooshtari, 2001; Zhao et al., 2020).

One of the essential determinants of perceived health status is the number of healthcare visits per year. According to one study, increasing the number of patient-provider counseling sessions will positively affect health-promoting lifestyles in patients experiencing health problems (Fallon et al., 2012). However, the evidence about the relationship between the number of healthcare visits and an individual’s perceived health status has been mixed. Based on the findings in one study, an increased number of healthcare visits is positively associated with promoting healthy behaviors, such as healthy eating habits in a group of people controlling for age, education, and income (Smith et al., 2017). The same study also found that higher healthcare visits within the past year are related to lower perceived health status among African American women with hypertension or obesity. This association was not modified significantly by covariates (age, income, and education) (Smith et al., 2017). In another study done in Finland, perceived health was inversely related to the number of physician contacts per year, and this was seen as consistent even when standardized by age, sex, and social status in a sample of adults in northeastern Finland (Miilunpalo et al., 1997).

Looking at the demographic characteristics of individuals can also give us valuable information about the higher risk of chance for some diseases in a particular group of people and the etiological factors in their disorders (Lubin et al., 1988). For instance, one study indicated that being Hispanic and age<45 is associated with less likelihood of receiving treatment for hypertension (Egan et al., 2014). While there is compelling evidence about the effect of sociodemographic characteristics on healthcare utilization (Ming et al., 2022; Zhou et al., 2020), it remains unclear how demographic factors modified the relationship between the number of healthcare visits and general health status.

This retrospective cross-sectional study examines how sociodemographic characteristics, including age, sex, race/ethnicity groups, education level, marital status, and annual household income, can change the association between an individual’s number of healthcare visits and their perceived health status. The result of this study will contribute to a better understanding of the need for interventions that address individuals’ barriers to having a better health status concerning their sociodemographic characteristics.

## Methods

### Study Design/Population

This retrospective cross-sectional study utilized data abstracted from the National Health and Nutrition Examination Survey (NHANES) for 2017-2018 (NCHS, 2017). NHANES is a population-based survey to assess the dietary intake, health, and nutritional status of noninstitutionalized adults and children in the United States. The data from NHANES are obtained through household interviews and standardized physical examinations. The NHANES interview includes demographic, socioeconomic, dietary, and health-related questions. The exposure of interest is the number of healthcare visits in the past 12 months, which was gathered using questionnaire data. The outcome of interest was perceived health status for the past 12 months, also from questionnaire data. Study participants meet the following inclusion criteria: ages 20-80 years old, complete data on perceived general health, race/ethnicity, education level, annual household income, gender, and number of healthcare visits in the past 12 months available. Sociodemographic factors were assessed by using data from questionnaires, as well as medical records and were categorized as follows: race (Non-Hispanic White, Non-Hispanic Black, Hispanic, other race), years of education (less than high school diploma, high school graduate/ GED, some college, college graduate or higher), and annual household income ($20,000 or less, $20,000-$44,999, $45,000-$74,999, $75,000-$99,999, $100,000 or more), gender (male or female), and marital status (married or not married).

### Statistical Analysis

A multiple logistic regression analysis was performed after using forward selection to choose sociodemographic factors, and the probability of entry (often denoted as α) was specified as p<0.05. Wald’s tests were used to determine the significance of covariates in the model. The dataset was cleaned for missing or incorrect data and recoded to minimize categories of each categorical variable. The model was evaluated for the goodness of fit using a Hosmer-Lemeshow test. It was determined that there is no evidence of a lack of fit in the model (p= 0.35). All analysis was performed in Stata statistical software.

## Results

The sample included 4,755 individual participants’ data. 1,181 (24.8%) participants had poor or fair health, and 3,574 (75.2%) had good or better health. The study population characteristics are summarized in **Table 1**. On average, participants with good or better health were more likely to be female (52.0% vs. 48.0%), younger (mean age of 50.2 vs. 55.5 years), White (28.5% vs. 8.3%), and more likely to be married (48.0% vs. 47.8%). Those with good or better health were also, on average, found to be in higher income groups and have more education (p<0.05). The variables included in the final model are reported in **Table 2**.

**Table 1.** Sociodemographic characteristics of the participants at baseline

| Baseline Characteristics | Perceived Health Status |  |
| --- | --- | --- |
|  | Poor/Fair Health<br>N=4,755 | Good/Better Health |
| Age | 1,181 (24.8%) | 3,574 (75.2%) |
| Race |  |  |
| Hispanic | 355 (7.5%) | 651 (13.7%) |
| Non-Hispanic White | 397 (8.3%) | 1,354 (28.5%) |
| Non-Hispanic Black | 257 (5.4%) | 825 (17.4%) |
| other | 172 (3.6%) | 744 (15.6%) |
| Gender |  |  |
| Male | 565 (11.9%) | 1,738 (36.6%) |
| Female | 616 (13%) | 1,836 (38.6%) |
| Education |  |  |
| Less than High School | 412 (8.7%) | 478 (10.1%) |
| High School Graduate or GED | 309 (6.5%) | 845 (17.8%) |
| Some College | 326 (6.9%) | 1,228 (25.8%) |
| College Graduate or Higher | 134 (2.8%) | 1,023 (21.5%) |
| Marital Status |  |  |
| Married | 665 (14%) | 1,714 (36%) |
| Not Married | 516 (10.9%) | 1,860 (39.1%) |
| Annual Household Income |  |  |
| \$20,000 or less | 382 (8%) | 546 (11.5%) |
| \$20,000-\$44,999 | 416 (8.7%) | 1,011 (21.3%) |
| \$45,000-\$74,999 | 199 (4.2%) | 766 (16.1%) |
| \$75,000-\$99,999 | 75 (1.6%) | 406 (8.5%) |
| \$100,000 or more | 109 (2.3%) | 845 (17.8%) |
| Number of Healthcare Visits |  |  |
| 0 visit | 152 (3.2%) | 619 (13%) |
| 1 visit | 108 (2.3%) | 714 (15%) |
| 2-3 visits | 246 (5.2%) | 1,141 (24%) |
| 4-5 visits | 226 (4.8%) | 525 (11%) |
| 5-6 visits | 111 (2.3%) | 205 (4.3%) |
| 7-8 visits | 62 (1.3%) | 87 (1.8%) |
| 9-10 visits | 118 (2.5%) | 142 (3%) |
| 11-12 visits | 50 (1.1%) | 48 (1%) |
| 13-14 visits | 108 (2.3%) | 93 (2%) |

**Table 2.**
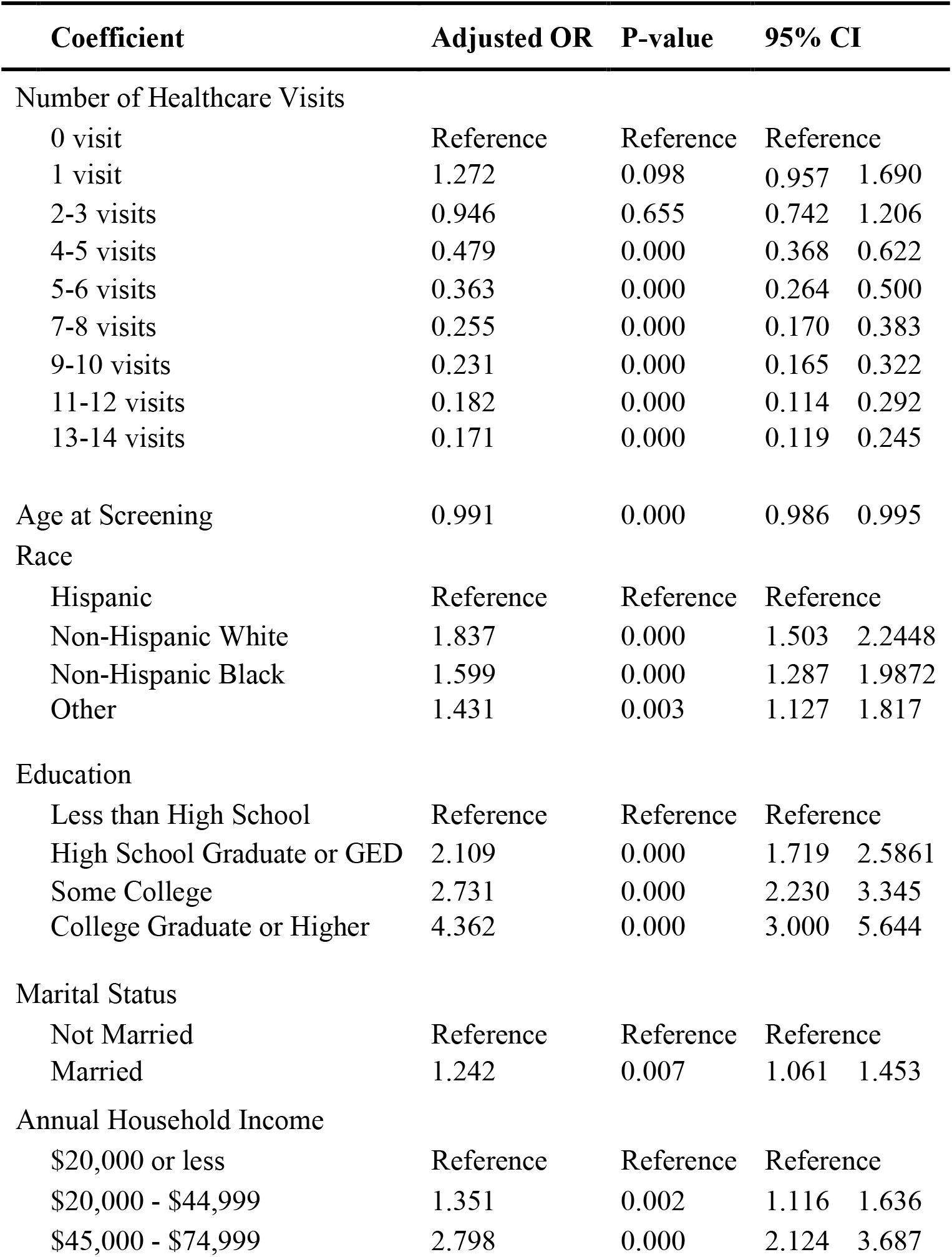
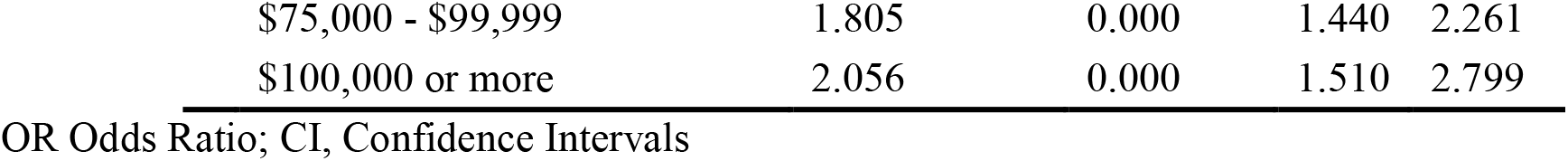
Odds ratios and confidence intervals from multiple logistic regression model of the number of healthcare visits as a function of perceived health status and adjusted covariates listed below

Results from the multiple logistic regression analyses are summarized in **Table 2**. Gender was not found to be a significant predictor of perceived general health using a forward model selection (p=0.64). All other predictors were found to predict perceived general health (p<0.05) significantly. When controlling for all significant covariates, participants who received 4-5 healthcare visits were less likely to report having poor or fair health (OR=0.48, p<0.001). The odds of good or better health consistently decreased as healthcare visits increased when adjusting for all other significant predictors.

## Discussion

This retrospective study examined the association between the number of healthcare visits in 12 months and perceived health status. Another purpose of this study was to determine if sociodemographic characteristics can modify this relationship between the reported number of healthcare visits and perceived health status.

According to the results of this study, there is a statistically significant association between the number of healthcare visits and perceived health status. As the number of individual healthcare visits increases in 12 months, the probability of having a good or better perception of health status decreases. This association was maintained after adjusting for sociodemographic characteristics. In line with the results of this study, a prior study found that the self-reported number of healthcare visits is significantly negatively associated with perceived health status (Smith et al., 2017).

Gender as a demographic feature did not significantly predict the relationship between the number of healthcare visits and perceived health. Other sociodemographic characteristics, including age, race, education, marital status, and annual household income, are all significant predictors in this model.

Our findings reveal notable racial disparities in perceived health within our sample study. Specifically, we observed that only 13.7% of Hispanic and 17.4% of Non-Hispanic Black individuals rated their health as good or better, compared to 28.5% of Non-Hispanic White individuals. Regarding the level of education, individuals with higher levels of education have a better perception of their general health. This finding was consistent with the previous study’s finding that low literacy is associated with self-reported poor health because health literacy is essential to healthcare access (Baker et al., 1997).

Finally, we found that the relationship between the number of healthcare visits within 12 months and perceived health status was mediated by income. Individuals with sufficient income were more likely to perceive their health status better. In support of our finding, one study revealed that low-income Black people have lower self-reported health (Franks et al., 2003).

This study will have several strengths and limitations worth highlighting. The large sample size in this study enables us to generalize the results to a wider range of populations, increasing the external validity. In addition, to reduce potential confounding in the study, we adjusted for several sociodemographic characteristics. Regarding the limitations of this study, using a self-report questionnaire for estimating perceived health status is not a reliable measurement of an individual’s general health condition since some participants might overestimate the times that they visited a healthcare provider within a 12-month. While we covered several sociodemographic factors that could affect a person’s perception of their health status, many other factors, including diet, exercise, and mental health, play an essential role in the relationship between the number of healthcare visits and perceived health status. Future research should be pursued to better understand the role of other factors related to perceived health status.

## Data Availability

All data produced are available online at: https://wwwn.cdc.gov/nchs/nhanes/continuousnhanes/overview.aspx?BeginYear=2017.

https://wwwn.cdc.gov/nchs/nhanes/search/datapage.aspx?Component=Questionnaire&Cycle=2017-2018

